# Qualification of the UniBw protection concept in different rooms of the Obermenzinger high school

**DOI:** 10.1101/2021.03.12.21253265

**Authors:** Christian J. Kähler, Thomas Fuchs, Rainer Hain

**Affiliations:** Universität der Bundeswehr München, Institute of Fluid Mechanics and Aerodynamics, Werner-Heisenberg-Weg 39, 85577 Neubiberg, Germany

## Abstract

Current research has shown that SARS-CoV-2 is transmitted via airborne particles. These particles are generated when infected persons exhale and they spread throughout the room, resulting in a high concentration and thus a high risk of infection of non-infected persons. One way to reduce the concentration of particles is to filter them using mobile room air cleaners, which can be easily set up and reduce the concentration of viruses or keep them at a low level. Since many studies are conducted in generic rooms, the question of the cleaning performance of such room air cleaners in real rooms arises. In order to investigate this, measurements of the effectiveness were carried out in a total of 4 different rooms of the “Obermenzinger Gymnasium” (Obermenzinger high school) in Munich. It can be shown that good effects of the room air cleaners are achieved even under realistic conditions. Even Plexiglas screens, which serve as protection against direct infection, have no negative influence.

## 1. Introduction

For many months, there has been a very controversial public debate on whether or not the protective walls and mobile air cleaners that have long been established in the adult world should also be installed in school classrooms as proposed in [1]. The German Physical Society recently clarified in an open letter that only mechanical ventilation can provide a high level of safety against indirect SARS-CoV-2 infection [2]. It has also pointed out the importance of protective walls to reduce direct infections and urged their installation in schools [3, 4]. The Harvard T.H. Chan School of Public Health [5] and Centers for Disease Control and Prevention (CDC) [6] are recommending mobile air cleaners and transparent protection walls to enhance the infection protection in schools. Transparent protective walls are already ubiquitous in many private sector work areas, as well as in public facilities and government offices, and many state parliaments and courtrooms. Mobile air cleaners with H13 / H14 class filters, which filter at least 6 times the room volume per hour, are used in many areas where adequate virus reduction cannot be achieved by heating, ventilation and air conditioning (HVAC) systems. In classrooms, on the other hand, the widespread introduction of these protective measures is prevented by those in power with changing arguments, with reference to the handout “Lüften an Schulen” (Ventilation in Schools) from the Federal Environment Agency (UBA) [7]. The background to this tactic can only be guessed, see https://www.news4teachers.de/2021/02/der-luftfilter-skandal-wie-bundesbildungsministerium-und-umweltbundesamt-den-einsatz-der-geraete-in-schulen-schlechtreden-und-was-dahintersteckt/.

The UBA only recommends opening all existing windows in the classrooms for 3−5 minutes widely every 20 minutes and for the entire length of the break. According to the UBA, the goal of the measure is to completely exchange the air 3 times per hour [7]. It has never been scientifically proven that this ventilation method really achieves the goal. There is also no evidence to what extent the UBA concept prevents infections in classrooms at all, because ventilation at best reduces the indirect risk of infection caused by a high viral load in the room. The much more dangerous direct risk of infection, which always exists when people are close together and the exhaled viral load can be inhaled directly by a neighboring person, as in the checkout room, cannot be prevented at all by ventilation. Why 3 air changes per hour should be sufficient is also not scientifically justified. It is claimed that 3 air changes with window ventilation should be equivalent to a 6-fold filtration of the room air. But this is wrong, because in both cases the reduction of viruses is based on mixed ventilation. Only if the cross ventilation is used, there would be a difference, because then the displacement ventilation prevails [8, 9]. But cross ventilation can only be achieved if a room has windows on both sides, which is very rarely the case. Opening a door is often considered as cross-ventilation, but this is wrong. According to the state of the art of ventilation technology, doors should not be opened either, because then the pollutants can get into other parts of the building in an uncontrolled way [8]. Due to the hazardous nature of the SARS-CoV-2 virus, air exchange rates of at least 6 or even 8 are required by other scientists [9, 10]. But also leading international institutions recommend air exchange rates larger 3 such as the Harvard T.H. Chan School of Public Health [5] and the CDC in the USA [6].

It is useful to be clear about what the UBA ventilation concept means. If a school lesson of 45 minutes duration is considered and it is assumed that ventilation takes place for 3−5 minutes after 20 minutes, this means that no ventilation takes place for almost 90% of the school lesson and thus the virus load in the room increases if one or more infected persons are in the room. How well the decrease in viral load occurs during intermittent ventilation depends on the number and size of windows, the temperature difference between indoors and outdoors, and the wind conditions outside the building [8, 9, 11]. Figure 1 illustrates the relationships. If there is no ventilation for more than one school hour, the viral load in the room increases linearly with time (red line). The increase in virus concentration, i.e. the number of viruses per cubic meter, depends on the strength of the source, i.e. the number of viruses released per unit time (*N*_*ssourse*_). In addition, the increase in virus concentration depends on the room size *V*_room_. The larger the space, the longer it takes to reach a certain virus load. Mathematically, this can be expressed by the following equation:

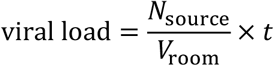

**Figure 1.**
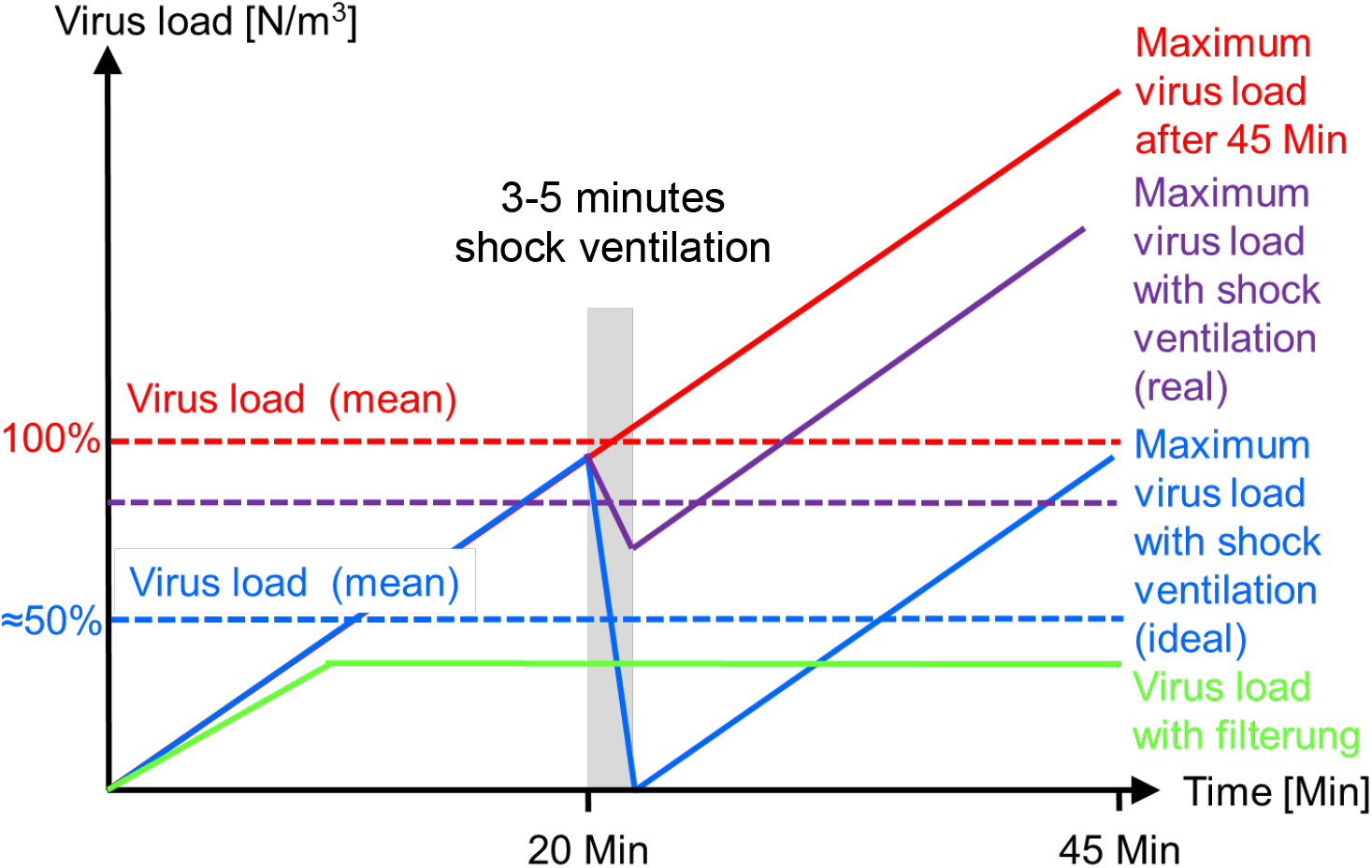
Increase of virus load in the room without measures (red), with ideal (blue) and real (purple) window ventilation and with mechanical filter technology (green) according to [12].

The number of viruses inhaled during the school day is important with regard to the probability of infection. This results from the integral over the respiratory flow rate multiplied by the viral load over the period under consideration. If the respiratory flow rate is assumed to be constant, this is equal to the respiratory flow rate multiplied by the mean viral load over the period under consideration. The red dashed line illustrates the mean value resulting from the corresponding curve.

If we now assume that after 20 minutes all windows in the classroom are opened for 3−5 minutes and all viruses leave the room through the window during this time, then the curve shown in blue (solid line) results. I.e. after the ventilation process the virus load increases again. The virus load over time can be reduced by a maximum of 50% in this way (blue dashed line). In practice, however, it will not be possible to remove all viruses from the room within 3−5 minutes [8, 9, 11]. As a rule, the decrease in virus concentration will be significantly less, as shown by the purple lines. This is partly due to the fact that not all windows are opened or are opened for too short a time, otherwise it becomes too cold in the room. Even if many windows are opened for a long period of time, temperature and wind conditions will often not allow 100% air exchange [8, 11]. The actual average viral load with shock ventilation will therefore always be between 50% and 100% of the reference case without ventilation.

The advantage of the technical separation of viruses with the help of mobile air cleaners is that with this method a continuous removal of the viruses from the room air takes place [13, 14, 15, 16, 17, 18, 19]. Furthermore, the separation of viruses is independent of whether people can or want to ventilate, and the number and size of windows and the temperature and wind conditions do not play a role either. Therefore, with appropriate airflow, the virus load can be maintained at a constant low level in the presence of sources, as illustrated by the green line in Figure 1.

It is often claimed that a major disadvantage of mobile air purifiers is their inability to remove CO_2_ from the room. Apart from the fact that there are mobile devices that can do this [15], the question arises as to why a device that is supposed to protect against potentially deadly viruses is required to also create a harmless gas from the room at the same time. Free ventilation is also not able to keep the temperature in the room on an acceptable level during winter time or to save energy and keep the children and youth from freezing in the room. It is also not expected from window ventilation to not interrupt lessons, or to ensure that fine dust, pollen and noise does not enter the room through the open windows.

The intensive debate in the media about the correct ventilation concept in schools has led to the assumption that a sufficiently rapid removal of the viruses from the room air is sufficient to ensure safety from a SARS-CoV-2 infection. However, it must be taken into account that free ventilation via windows, HVAC systems and mobile air cleaners can only reduce the indirect risk of infection, i.e. infection due to the high viral load in the room. Ventilation of any kind cannot protect against the direct risk of infection, i.e. the direct uptake of viruses due to short distances to infected persons or insufficient protection by a particle-filtering FFP2/FFP3 mask [20] or protective walls. Since sufficient spacing in classrooms is often not feasible (with the possible exception of rotating classes) and FFP2/FFP3 masks should not be worn permanently, a study was conducted to determine whether transparent protective walls with a surrounding edge impair the filtering effect in classrooms [1]. The result of the study clarifies that the removal of aerosol particles from the room air does not depend on whether the room is equipped with tables and chairs, additional people with bags and laptops, and protective walls between people. This is physically understandable, since window ventilation and the use of mobile air cleaners is based on mixed ventilation. That is, the natural flow movements in the room, caused by the movement, breathing and heat emission of people, temperature differences in the different areas of the room and flow movements due to open windows or mobile air cleaners, provide constant mixing of air masses [8, 11]. For this reason, all areas of the room are also quite evenly cleared of the viral load, regardless of where the windows or mobile air cleaners are positioned [13, 17]. The greater the mixing, the better the cleaning performance in distant areas. Therefore, any concerns that people walking around may reduce filter performance are physically incorrect. It is also false that protective walls between seat neighbors at a table would affect classroom ventilation, as insinuated in [21]. These insinuations are based on a misconception of mixed ventilation and unvalidated simulation results that cannot correctly reproduce such situations [13, 17, 22].

The protection concept presented and experimentally validated in [1] is increasingly being established in schools and there are now entire regions and cities in Germany that rely on this protection concept to protect children and young people. With regard to the Swiss cheese model of the pandemic [23], this implementation of the protection concept is to be considered very useful, as it represents an effective building block for the prevention of infections. Especially with regard to future virus variants whose spread is more difficult to control due to mutations, the use of further protective measures is imperative to prevent an exponential growth of infection numbers. Therefore, it is no wonder that the German Association of Physics [2, 3], the Harvard T.H. Chan School of Public Health [5] and Centers for Disease Control and Prevention [6] are recommending mobile air cleaners and transparent protection walls to enhance the infection protection in schools. The question is often asked how effective the protection concept presented in [1] is in different rooms of a school. To answer this question, experiments were conducted in the Obermenzing high school (Obermenzinger Gymnasium). In total, four rooms with different geometry were analyzed. In the following, the experiments and the results are presented and discussed.

## 2. Measurement setup and data analysis

Since the effectiveness of the protection concept in normal classrooms with a rectangular floor plan and typical table arrangement has already been proven [1], a computer room with U-shaped table placement against the wall (room A), a teachers’ room with distributed tables (room B), a classroom with a gable roof (room C) and a canteen with an attached kitchen (room D) were analyzed in this study. Figures 2–5 schematically show the top view of the rooms with the room equipment and the position of the measuring device and the room air cleaner. The distance between the measuring device and the room air cleaner was chosen as large as possible in all cases in order to analyze the most unfavorable conditions in each case.

**Figure 2:**
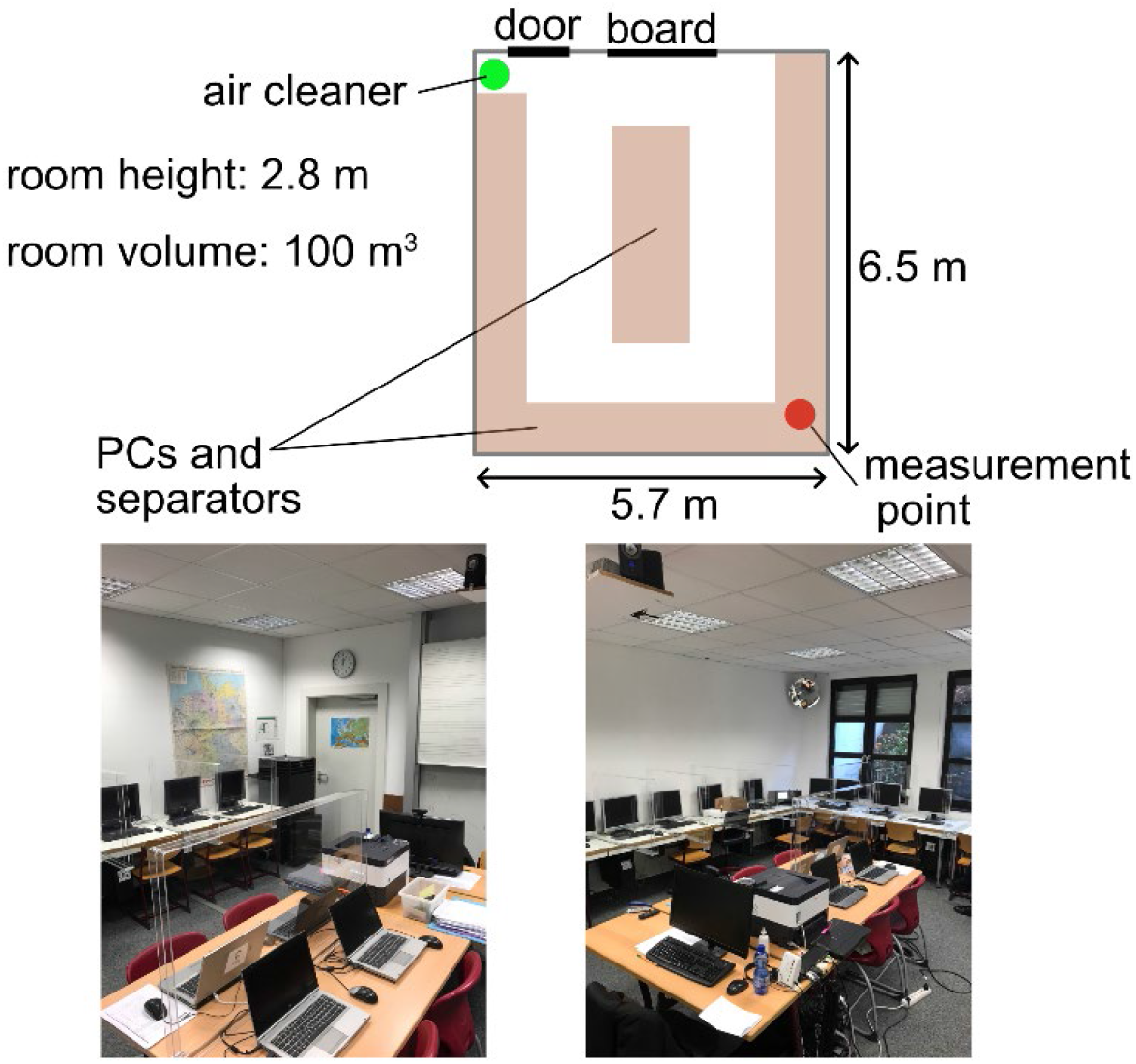
Room A – Computer room.

**Figure 3:**
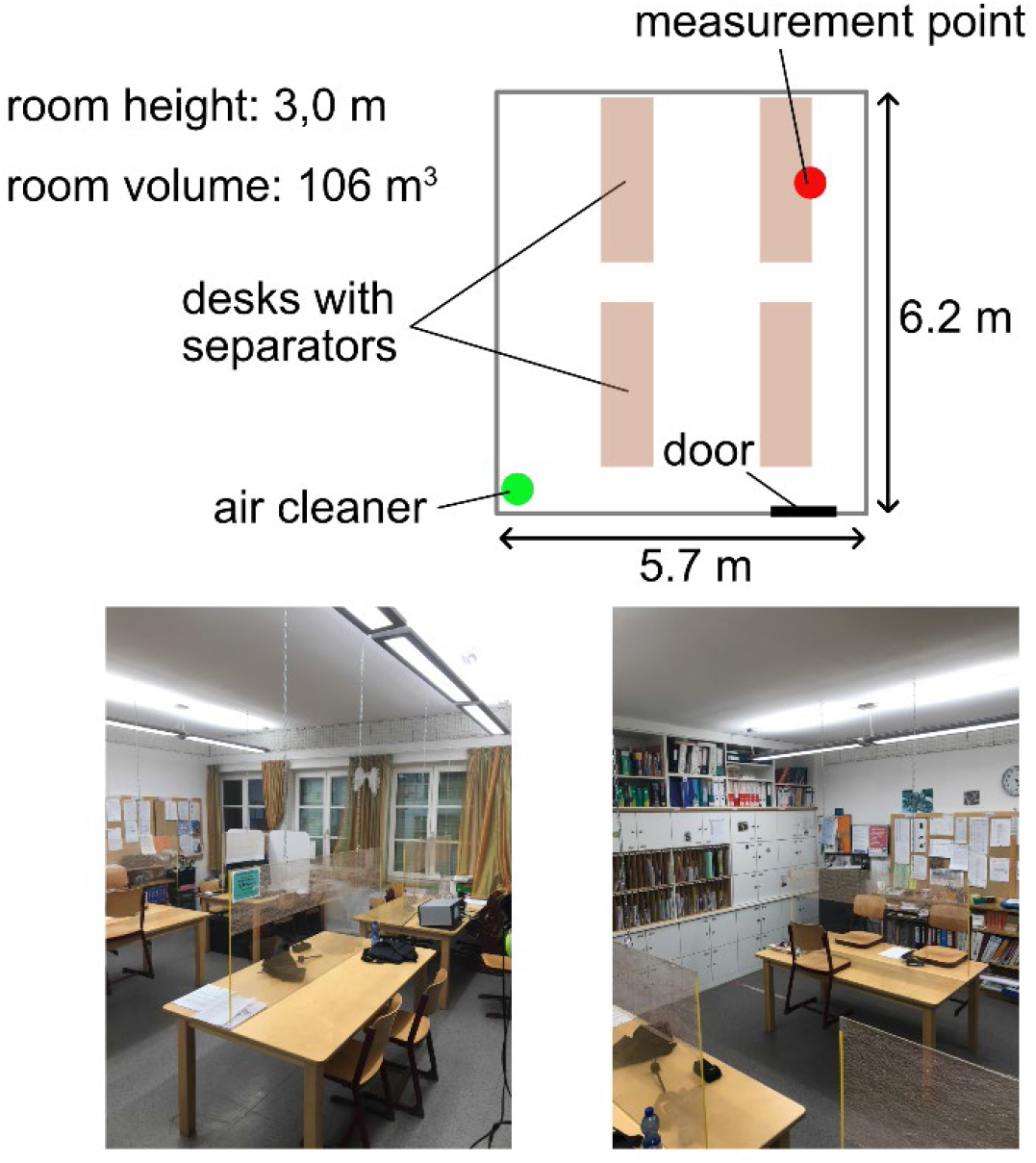
Room B – Teachers’ room.

**Figure 4:**
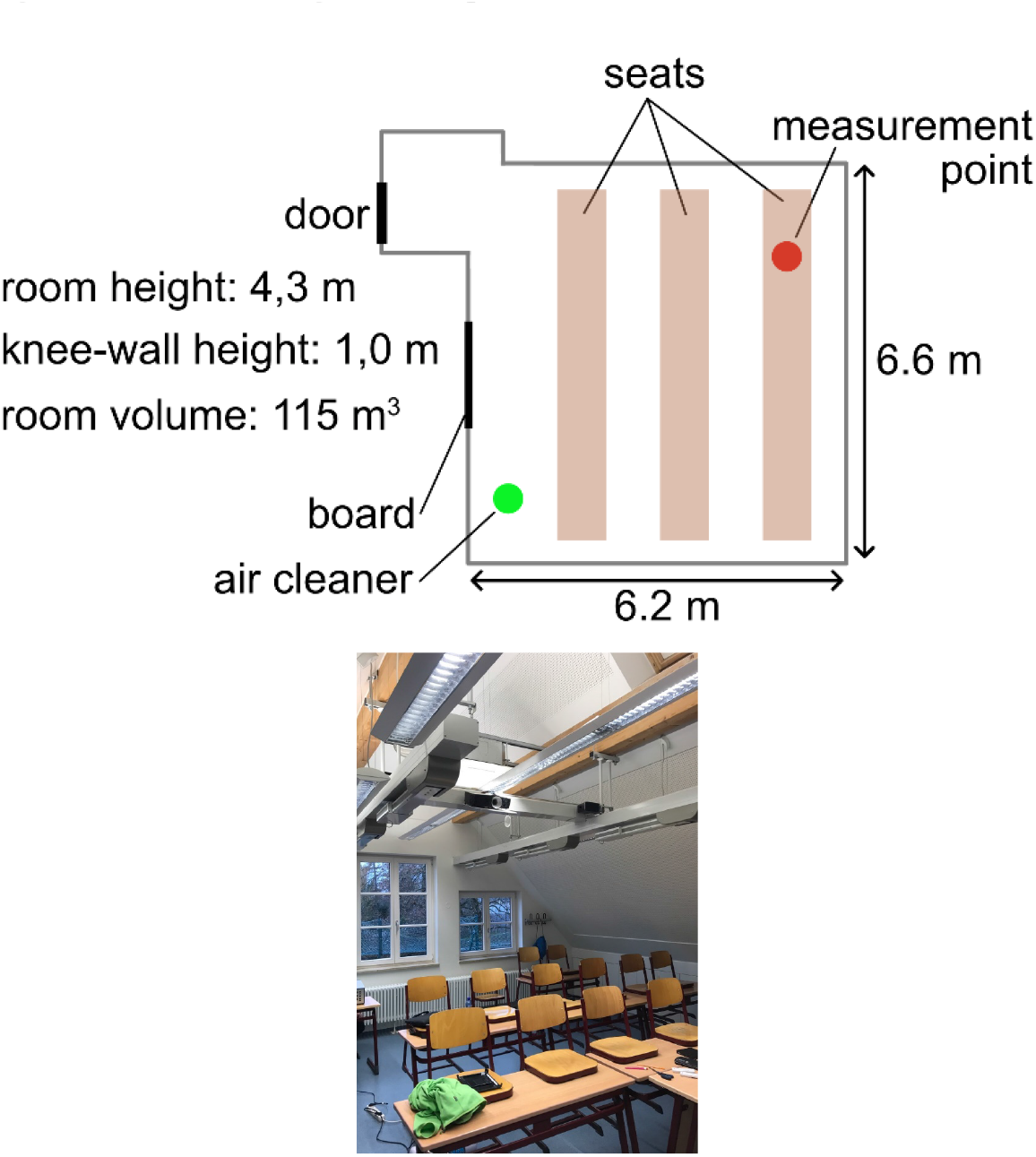
Room C – Classroom with gable roof.

**Figure 5:**
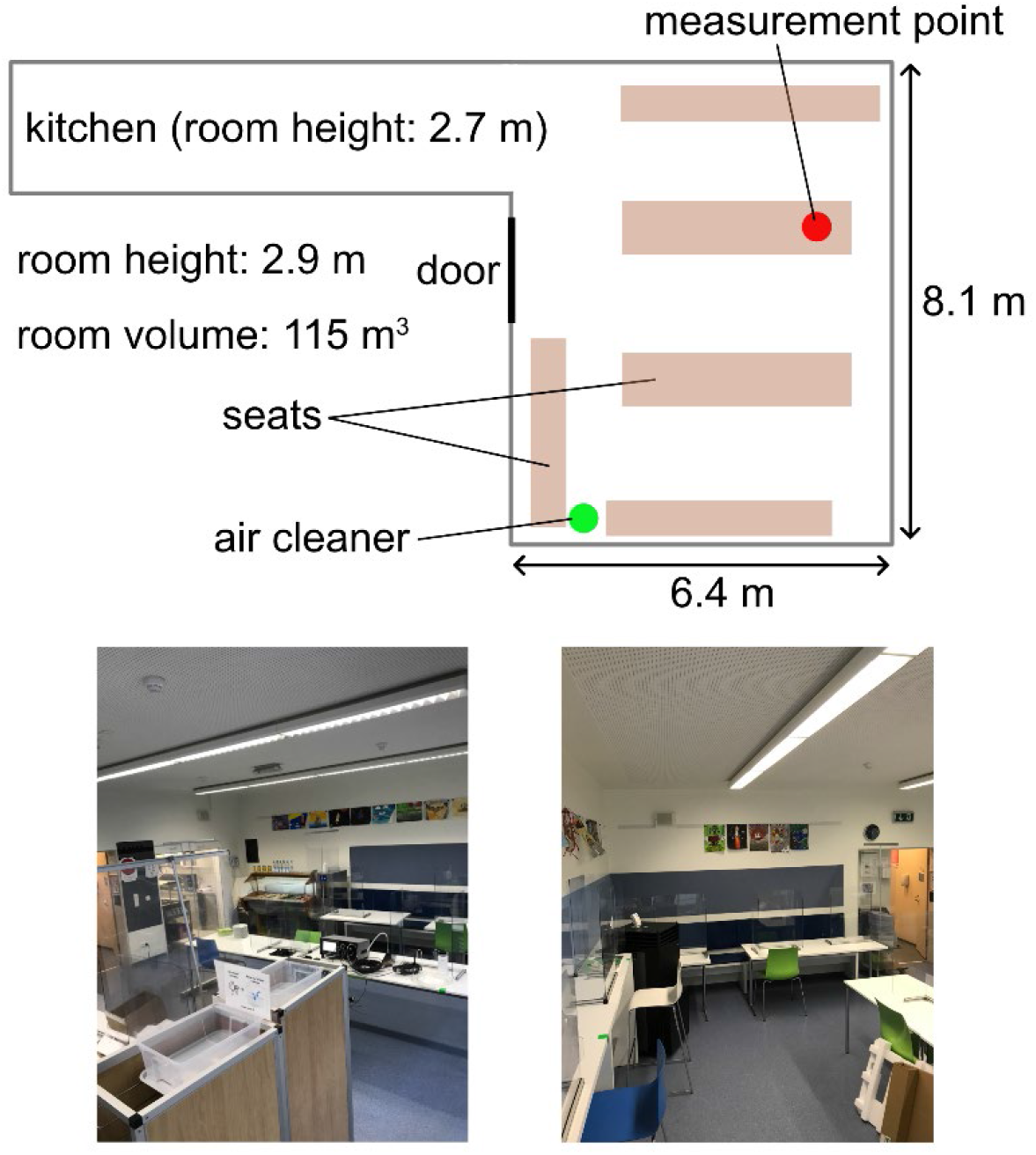
Room D – Canteen.

For the determination of the cleaning efficiency, artificially generated aerosol particles of DEHS (average diameter approx. 0.4 µm) were introduced into the respective room and homogeneously distributed before the start of the measurement. The size of these particles is in the range of aerosol particles emitted by infected humans and contaminated with viruses [24, 25, 26]. The particles follow the flow in the room almost ideally and do not exhibit any noticeable evaporation during the period under consideration. The filtering of the aerosol particles was realized with a TROTEC TAC V+. The volume flow rate was set in such a way that at least a theoretical air exchange rate of *k*_theo_ > 6 was obtained (*k*_theo_ = 8 in room A; *k*_theo_ = 7.5 in room B; *k*_theo_ = 7 in room C; *k*_theo_ = 8.7 in room D). The theoretical air exchange rate results from the ratio of the room air cleaner volume flow to the room volume. The time course of the particle concentration was recorded using a *Promo 3000* particle counter from Palas GmbH (Germany) with a *Welas 2300* sensor head.

## 3. Measurement results

The measured, normalized particle concentrations over time are shown in Figure 6. Before each measurement with the air cleaner running, reference measurements were first carried out with the air cleaner switched off. This is necessary to determine how many aerosol particles leave the room through leaks or settle on the walls. The respective course shows that rooms A – C are altogether quite dense. Room D, on the other hand, already shows quite a large decrease in aerosol particles without an air purifier. This decrease is caused by the operation of exhaust air systems in the kitchen and an open door in the dining area to the hallway. This configuration was chosen for the measurement because it corresponds exactly to the configuration that exists in real school operation. When the air cleaner is operated, there is a fairly rapid decrease in the aerosol particle concentration in all rooms. So when these systems exist or HVAC systems, they should be used. However, it should be ensured that the removed air is replaced by fresh air from outside and not by contaminated air from other rooms or floors.

**Figure 6:**
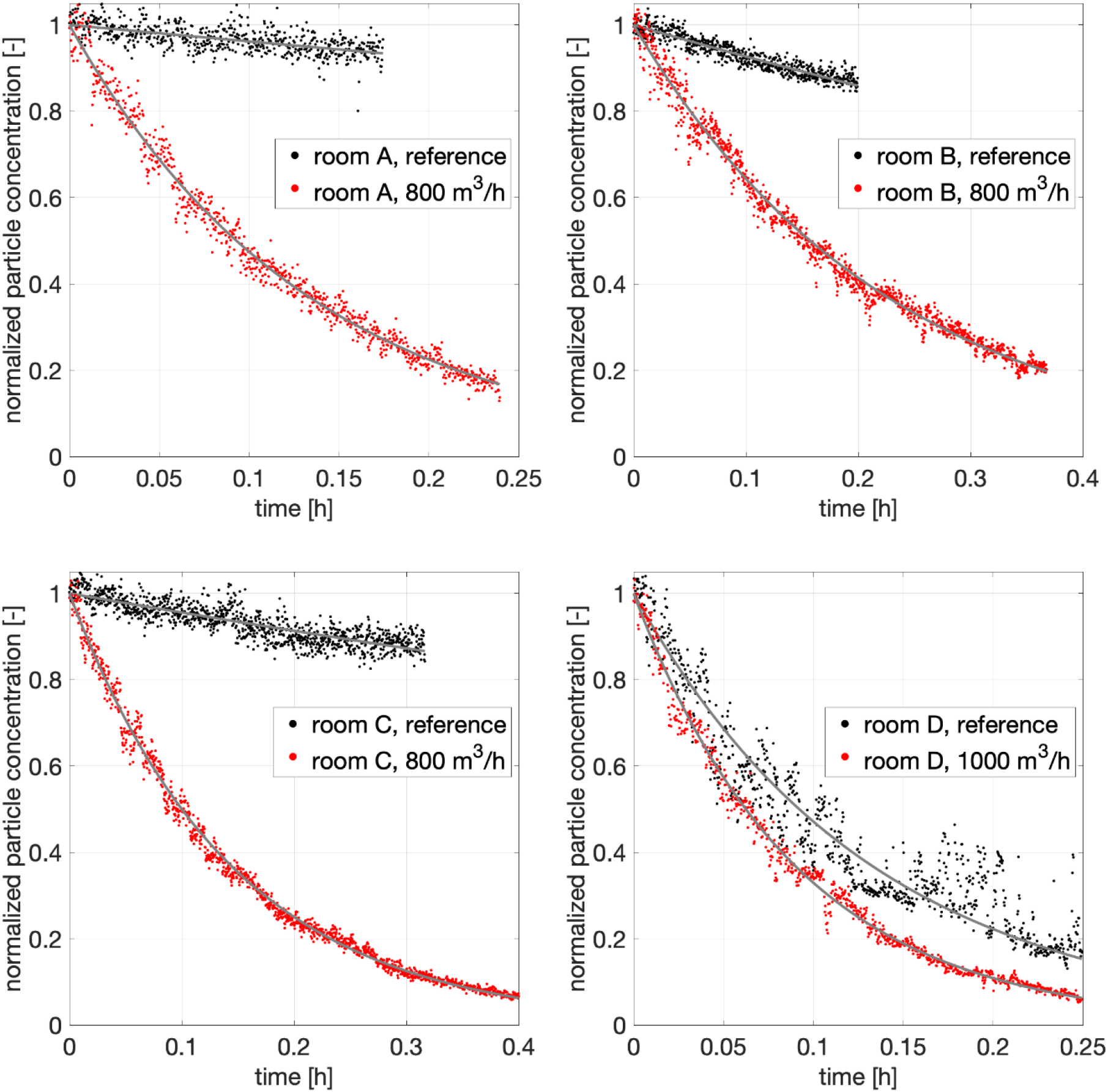
Normalized particle concentrations vs. time for the 4 measured rooms.

The decay rate *k* with the unit [1/h] is determined from the measured particle concentrations over time. In ventilation technology, this is also known as air exchange rate, air exchange rate or air exchange rate. With the aid of *k*, the development of the particle concentration *c* over time can be determined, provided that the room volume *V* [m^3^] and the strength *S* [particles/h] of the source of contamination are known. The steady-state concentration *c*_steady_, that occurs after a long time as indicated in figure 1 can be calculated as follows:

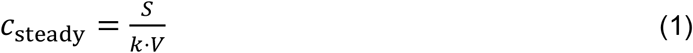

The higher *k* is, the faster potentially hazardous aerosol particles are removed, or the lower the concentration will be after a longer period of time. The decay rates determined for the different rooms are shown in Table 1.

**Table 1:**
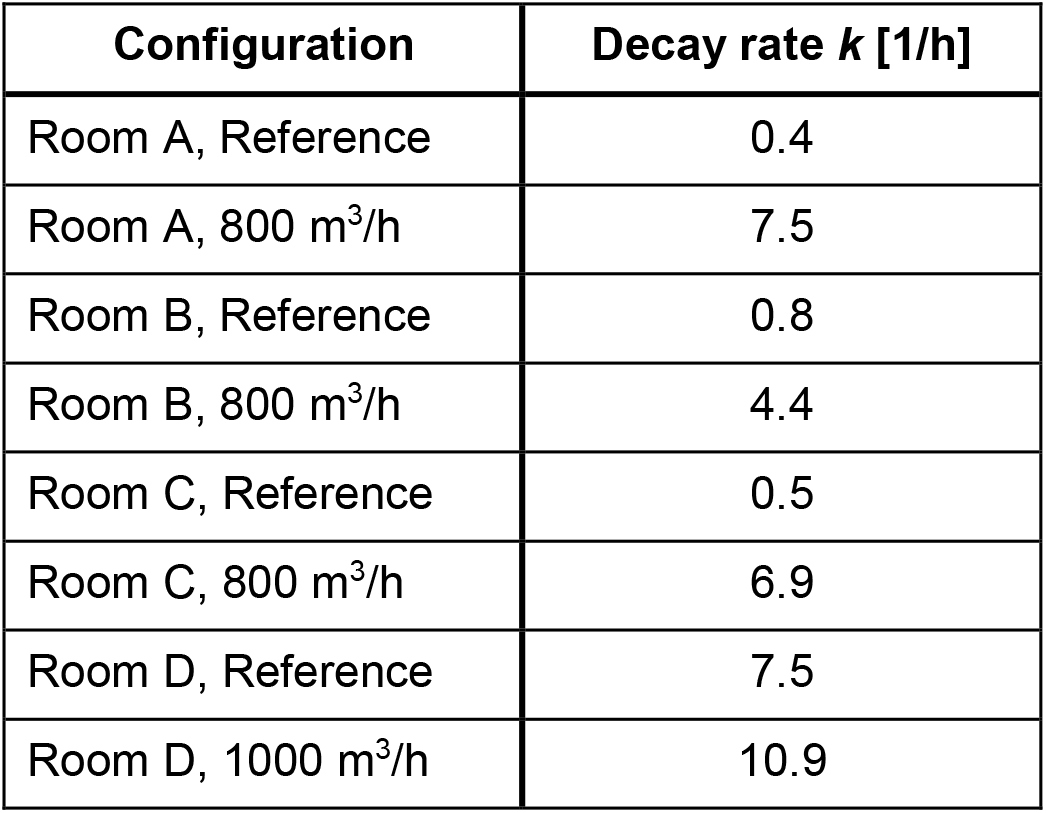
Decay rates determined for different configurations.

| Configuration | Decay rate $k$ [1/h] |
| --- | --- |
| Room A, Reference | 0.4 |
| Room A, 800 m<sup>3</sup>/h | 7.5 |
| Room B, Reference | 0.8 |
| Room B, 800 m<sup>3</sup>/h | 4.4 |
| Room C, Reference | 0.5 |
| Room C, 800 m<sup>3</sup>/h | 6.9 |
| Room D, Reference | 7.5 |
| Room D, 1000 m<sup>3</sup>/h | 10.9 |

It can be seen that for rooms A and C there is good agreement between the measured air exchange rate and the theoretical air exchange rate. In room A, the measuring sensor was located in the diagonally opposite corner of the room directly between two transparent protective walls. Even in this unfavorable position, the measured filter efficiency agrees well with the theoretical value. Therefore, it is shown that the transparent protective walls with surrounding edges have no negative effect on the filter performance, as claimed in [21] without any reliable evidence. This also means that window ventilation in combination with the transparent protective walls is not negatively affected. Physically, the result is clear, since in both cases it is a mixed ventilation, as already stated at the beginning. In room B, *k* was lower than *k*_theo_. The reason for this is an open door to the corridor. This result shows that the entire room volume must always be taken into account when determining the correct volume flow. This means that if adjacent rooms are not separated by closed doors, then the volume of the adjacent room must be taken into account when setting the volume flow, since the room air in the adjacent room is also captured by the filter unit. From this corridor, the entrance door leads directly to the outside. This door was kept closed for the measurements, but in practical operation it is frequently opened and another door in the corridor is not always closed in real operation as in the tests carried out here. This results in additional ventilation in real operation, which increases *k* further. The result in room D shows that, in addition to the air cleaner, other ventilation measures can be used to accelerate the reduction of the viral load in a room if necessary. However, the increase in the *k*-value during operation of the room air purifier is not as large as theoretically expected. Presumably, this is due to the fact that the already high *k*-values combined with the flow through the room reduce the efficiency of the mixed ventilation. This is a typical phenomena that applies to window ventilation as well as mobile filter systems and other ventilation systems based on mixed ventilation [9, 15]. Nevertheless, even in this case the cleaning is supported and thus it is possible to supplement an ineffective window ventilation or too weak HVAC system with the help of mobile room air cleaners or to replace the window ventilation completely with a technical solution.

## 4. Conclusion

The results of the tests show that good filter performance can be achieved with mobile air cleaners in very different room situations. The placement of tables and chairs, computers and transparent protective walls in the room has no adverse effect on filter performance. Therefore, it is possible to reduce both the indirect infection risk in rooms with the mobile air purifiers and at the same time the much more dangerous direct infection risk by means of transparent protective walls with a surrounding edge. In order to counteract an increase in CO_2_ concentration in the room, the windows can be opened from time to time as in the past. This does not hinder the filter effect and the transparent protective walls also have no negative effect on CO_2_ exchange, as the mixed ventilation is always effective. Alternatively, room air cleaners can be used that filter the room air in recirculation mode and simultaneously feed outside air into the room via a bypass. In this way, the virus and CO_2_ problem can be solved technically at the same time, without having to constantly interrupt classes for ventilation. An important result of the study is also that inefficient ventilation through windows, doors or exhaust air systems can always be improved by mobile room air cleaners. However, powerful room air cleaners are able to keep the virus load in a room at a low level, or to quickly reduce a high virus load, even without window ventilation, because, in contrast to intermittent ventilation, they continuously ensure a reduction in the virus load and because the filter performance is completely independent of whether there are sufficient windows in the rooms, people are willing to ventilate, or the temperature and wind conditions can physically enable an air exchange at all. Therefore, regardless of the number and size of windows, people’s willingness or ability to ventilate, and physics, powerful mobile air purifiers with H13 or H14 class filters ensure consistent separation of the virus load in the room. It is therefore completely incomprehensible why the Federal Environment Agency is fighting the implementation of the scientifically proven protection concept. It is certainly not one of the tasks of the Federal Environment Agency to prevent effective protection concepts. They should accept the basics of fluid physics and follow the recommendations of institutions such as the DPG or CDC to protect children in schools during the pandemic and after.

### Note

The investigations were financially supported by the company TROTEC GmbH (Heinsberg, Germany). The investigations were carried out in compliance with the good scientific practice of the German Research Foundation (DFG). The support by the company TROTEC has no effect on the results presented.

## Data Availability

Data can be provided upon request.

